# Global Assessment of Palliative Care Need: Serious Health-Related Suffering Measurement Methodology

**DOI:** 10.1101/2024.02.26.24303409

**Authors:** Xiaoxiao J Kwete, Afsan Bhadelia, Héctor Arreola-Ornelas, Oscar Mendez, William E. Rosa, Stephen Connor, Julia Downing, Dean Jamison, David Watkins, Renzo Calderon, Jim Cleary, Joe Friedman, Liliana De Lima, Christian Ntizimira, Tania Pastrana, Pedro E. Pérez-Cruz, Dingle Spence, M.R. Rajagopal, Valentina Vargas Enciso, Eric L. Krakauer, Lukas Radbruch, Felicia Marie Knaul

**Affiliations:** University of Miami Institute for Advanced Study of the Americas, University of Miami, Miami, FL, USA; Yangzhou Philosophy and Social Science Research and Communication Center, Yangzhou, China; Department of Public Health, College of Health and Human Sciences, Purdue University, West Lafayette, IN, USA; Institute for Obesity Research & School of Government and Public Transformation, Tecnológico de Monterrey, Monterrey, Mexico; Tómatelo a Pecho, A.C., Mexico City, Mexico; Fundación Mexicana para la Salud (FUNSALUD), Mexico City, México; Department of Psychiatry and Behavioral Sciences, Memorial Sloan Kettering Cancer Center, New York, NY, USA; Worldwide Hospice Palliative Care Alliance, London, UK; International Children’s Palliative Care Network, Bristol, UK; University of California, San Francisco, USA; Department of Global Health, University of Washington, Seattle, WA, USA; Indiana University School of Medicine, Indianapolis, IN, USA; University of California in Los Angelas, School of Medicine, Los Angeles, CA, USA; International Association of Hospice and Palliative Care, Houston, TX, USA; African Center for Research on End of Life Care, Kigali, Rwanda; Department for Palliative Medicine, RWTH Aachen University, Germany; Programa Medicina Paliativa y Cuidados Continuos, Facultad de Medicina, Pontificia Universidad Católica de Chile (PUC), Santiago, Chile; University of the West Indies, Mona, Jamaica; Pallium India Trust, Kerala, India; Department of Global Health & Social Medicine, Harvard Medical School, Boston, MA, USA; University of Bonn, Bonn, Germany; Miller School of Medicine, University of Miami, USA; Leonard M. Miller School of Medicine, University of Miami, Miami, Florida, USA

**Keywords:** serious health-related suffering, palliative care, suffering measurement, palliative care need

## Abstract

Inequities and gaps in palliative care access are a serious impediment to health systems especially low- and middle-income countries and the accurate measurement of need across health conditions is a critical step to understanding and addressing the issue. Serious Health-related Suffering (SHS) is a novel methodology to measure the palliative care need and was originally developed by The Lancet Commission on Global Access to Palliative Care and Pain Relief. In 2015, the first iteration – SHS 1.0 – was estimated at over 61 million people worldwide experiencing at least 6 billion days of SHS annually as a result of life-limiting and life-threatening conditions. In this paper, an updated methodology - SHS2.0 - is presented building on the work of the Lancet Commission and detailing calculations, data requirements, limitations, and assumptions. The updates to the original methodology focus on measuring the number of people who die with (decedents) or live with (non-decedents) SHS in a given year to assess the number of people in need of palliative care across health conditions and populations. Detail on the methodology for measuring the number of days of SHS that was pioneered by the Lancet Commission, is also shared, as this second measure is essential for determining the health system responses that are necessary to address palliative care need and must be a priority for future methodological work on SHS. The discussion encompasses opportunities for applying SHS to future policy making assessment of future research priorities particularly in light of the dearth of data from low- and middle-income countries, and sharing of directions for future work to develop SHS 3.0.

## I. Background

Over 60 million people annually experience serious health-related suffering (SHS) that is amenable to palliative care. However, most reside in low-resource and rerual areas with nonexistent or inadequate palliative care services, and limited access to medicines and technologies that can reduce SHS,(1) emblematic of the tragedy and injustice of overall disparities in healthcare. Palliative care is a core component of universal health coverage (UHC), making the lack of access to palliative care a serious impediment to SDG Goal 3, namely, to “ensure healthy lives and promote well-being for all at all ages”(2, 3) and to achieving Sustainable Development Goal 10 focused on reducing inequality within and among all countries.(1, 2)

Efforts to address this global health failing and to close the divide in access to palliative care have been thwarted by various factors.(1, 4) One is the dearth of methods and data to quantify global palliative care need and this was a major area of work of The Lancet Commission on Global Access to Palliative Care and Pain Relief (hereafter referred to as Lancet Commission or the Commission) in developing SHS. Although evidence is required to develop appropriate and targeted recommendations for closing gaps in access to palliative care, measurement of the burden of SHS has not kept pace with progress in measuring the burden of disease.(1, 5) A scientific focus on measurement of SHS(6, 7) is a necessary complement to existing measures of the burden of disease such as quality-adjusted life years (QALYs) and disability adjusted life years (DALYs). Further, measurement of SHS has value and purpose in its own right as a global health issue and as part of efforts to achieve the SDGs.

The Lancet Commission report presented a breakthrough by introducing the concept of serious health-related suffering (SHS) to quantify the global and country-specific need for palliative care and pain relief in terms of both the number of individuals who experience SHS (population in need of palliative care services), and the number of days of each type of SHS (as an input to develop more effective health system responses to address palliative care need) in a given year. Building on more limited efforts to measure population-based need for palliative care in previous publications,(4) the Commission estimated the 2015 global burden of SHS at 61 million: 25.5 million people who died - 45% of the 56.2 million deaths worldwide - and an additional 35.5 million people who experienced an SHS-associated condition and did not die in that year, with at least 6 billion symptom days experienced by those people. The estimates were calculated by a systematic process documented briefly in the Lancet Commission report and in its entirety in an white paper.(1, 8)

The Lancet Commission Report has been cited by over 1000 research article publications as of this writing, and the data has been used by various international organizations and initiatives including the International Narcotic Control Board (INCB), the Worldwide Hospice Palliative Care Alliance (WHPCA), and the Disease Control Priorities (3rd edition), as well as various country champions of palliative care in their evidence generation, policy making and advocacy endeavors.(9–11) The Lancet Commission Secretariat was transformed into an interdisciplinary Research Hub on Global Access to Palliative Care and Pain Relief - jointly led by the University of Miami Institute for Advanced Study of the Americas and the International Association for Hospice and Palliative Care - to promote evidence generation, dissemination, and translation to policy and practice to achieve universal access to palliative care. The research hub built on the original Commission methodology – SHS1.0 - to generate the next iteration – SHS 2.0.

In this paper, the SHS 2.0 methodology is summarized, exclusively dealing with measuring the number of people who die with (decedents) or live with (non-decedents) SHS. The assumptions, strengths, and weaknesses of both the original and the 2.0 iteration for measuring people with SHS are discussed. The methodology for measuring the number of days of SHS is also detailed. Pioneered by the Lancet Commission, measuring days with SHS is essential for determining the health system responses to palliative care need and although not undertaken as part of SHS2.0, must be a priority for future methodological work on SHS. A guide to calculating the burden of SHS is provided, including specific instructions on measuring the number of people who die with (decedents) or live with (non-decedents) SHS and the number of symptom days they experience annually, as well as secondary indicators that may be constructed with the SHS database. The paper concludes with a discussion on the potential applications of SHS data for researchers, policymakers, and practitioners as well as directions for future work and priorities for developing SHS 3.0. It is linked to another methods paper on measuring distributed opioid morphine equivalent (DOME) and comparing DOME against the need for palliative care (SHS).

## II. Defining and measuring SHS

Serious health-related suffering, as defined by the Lancet Commission, is the “pain, suffering, and severe distress associated with life-threatening or life-limiting health conditions and with end of life”(1) that cannot be relieved without medical intervention and that is potentially amenable to relief through palliative care. SHS is not bound by time or prognosis and includes complex, chronic or acute, life threatening or life-limiting health conditions.(12)

The definition of palliative care adopted by the Lancet Commission is the one used by the World Health Organization (WHO) at the time: “an approach that improves the quality of life of patients and their families facing the problems associated with life-threatening illness through the prevention and relief of suffering by means of early identification and assessment and treatment of pain and other problems, physical, psychosocial, and spiritual”.(1, 13) SHS 2.0 adopts the consensus-based definition spearheaded by the IAHPC that was initiated as one of the recommendations of the Commission report and engaged a group of global stakeholders from low, middle, and high-income countries. Specifically, “palliative care is the active holistic care of individuals across all ages with serious health-related suffering due to severe illness and especially of those near the end of life. It aims to improve the quality of life of patients, their families, and their caregivers.”(12)

The SHS burden is presented both as the number of people experiencing SHS due to life-limiting or life-threatening conditions and as the number of symptom-days of SHS experienced. Individuals experiencing SHS are distinguished as either decedents or non-decedents and the conditions, multipliers, and estimates in each differ. Decedents are defined as individuals who died within the year of calculation and are thus captured in the mortality database. Non-decedents are individuals who did not die within the year of calculation and are thus captured in the prevalence database. Non-decedent categories of SHS include conditions 1) that may have been cured but from which SHS persists; 2) from which patients recover but that nonetheless caused SHS; 3) with survival with chronic severe disability and with SHS symptoms; and 4) have a slowly progressive course. Symptom-days are defined as the number of days decedents and non-decedents lived with any symptoms and are calculated for each symptom and aggregated to measure total palliative care need. The latter is key to analyzing the response to SHS, for example in DOME for specific symptoms such as pain or dyspnea.

### General considerations in the selection processes

The selection of conditions, development of multipliers, and calculation of the number of people and days of SHS was informed by a literature search, individual and group expert discussions, and Delphi processes with online surveys for SHS 1.0 as described in the Appendix to the Commission report. Expert panel(s) of palliative care clinicians with experience providing clinical care in different parts of the world, especially in LMICs were engaged in the process.

To estimate symptoms and symptom duration (days of SHS), as part of the work of the Commission and SHS1.0, experts were asked to consider a typical patient with each of the conditions and based on their daily experience, to generate an estimate of the prevalence and duration of each symptom. During the expert consultation stage, including focus group discussions and semi-structured interviews, results from the literature review were presented. Experts were asked to provide responses and feedback based on their work experiences even when those experiences were contrary to the evidence presented to them. Either individually or in groups, all data and estimates were vetted, considering assumptions and limitations or gaps to ensure that all relevant aspects or scenarios are reasonably accounted for when possible. It is expected that these data will serve to provide content validity for estimation of the global burden of remediable suffering.(14) See appendix table 1 for a full list of the experts’ consensus building practices undertaken by the LC.

Finally, the Delphi method for consensus-building also was used to determine the duration (average number of days requiring palliative care) for which palliative care was needed for each of the conditions included in the database.(15) Experts were purposively sampled and were considered to be ‘informed individuals’(16) and ‘specialists’(17) within the field of palliative care, in this case palliative care.(18) Both rounds of the Delphi requested 18 palliative care experts living in LMICs to estimate the number of days of palliative care that would be required for a patient with each of the given conditions. The responses from the first round were pooled to identify a group average range and standard deviation for each condition. The second round of the Delphi presented respondents with the average range of days of palliative care with confidence intervals for each parameter. Experts were asked to respond again to the same questions based on knowledge of the group’s prior responses. The response rate for round one was 83% and for round two was 27%. Results from each round are presented in Tables 2. See appendix table 2 for the results from the rounds 1 and 2 of the Delphi study.Due to limited resources, estimation for symptom-days is only available from the Lancet Commission (SHS1.0) and was not updated for SHS 2.0.

#### Taking children in account in SHS 2.0

The initial SHS database from the Commission work did not differentiate the SHS burden experienced by adults and children. Hence for SHS 2.0 and in collaboration with and under the leadership of the International Childreńs Palliative Care Network (ICPCN) with the engagement of IAHPC and WHPCA, an additional expert panel was convened for SHS 2.0 comprised of 8 pediatric palliative care specialists from both high-income and low- and middle-income settings around the world. Literature review and analysis,(19) an online survey, two virtual meetings each lasting at least 90 minutes, and various internal discussions were conducted to differentiate the calculation of palliative care needs for children and adults in select conditions.

#### Time-series analysis

A major improvement for SHS 2.0 is the time-series analysis to incorporate the sensitivity of SHS to changes in disease trajectories, changes in pathogens, emergence of new diseases, and with the evolution of and advancements in medical technologies to address the burden of disease, each of which impacts the SHS burden. This gap was identified through the incorporation of time series mortality and prevalence data to analyze historical trends in the SHS burden. Data for 1990, 2000, 2010 and 2019 are presented in the updated calculations. Those years were selected to represent the earliest obtainable evidence, and data points every 10 years, and 2019 was selected as the most recent year since it was the most updated year of data at the time of the commencement of this analysis. The need to account for endogenous variables was particularly evident for people living with human immunodeficiency viruses (PLWHIV), as well as patients living with tuberculosis, cancer, or cerebrovascular disease, and for children.

#### Switching from WHO’s Global Health Estimates (GHE) to IHME’s Global Burden of Diseases (GBD) database

The Lancet Commission estimated the SHS burden in the most recent year of available data at the time (2015) and using WHO’s global mortality database, Global Health Estimates (GHE). However, due to the lack of prevalence data in GHE, non-decedents were computed using fixed survivor-to-deaths ratios generated from global disease-specific reports. This assumed that all countries experience the global average survivor-to-deaths ratio for all conditions with non-decedents categories, not accounting for country-level variation in the epidemiological profile of survivors and limiting the applicability of country-specific analyses.

SHS 2.0 was improved along several dimensions by using the GBD database released by the Institute for Health Metrics and Evaluation (IHME). Firstly, the GBD includes country-specific data on mortality and prevalence. The prevalence data strengthens the calculation of non-decedents with SHS. In addition, GBD data date back to 1990, permitting the calculation of the burden of SHS over three decades. Further, the Lancet Commission report defined children as being 0-15 years of age as more disaggregated data was not available. For SHS 2.0, children are defined as 0–19-year-olds to be consistent with other key publications on children’s palliative care need around the world using the GBD data break-down of age groups.(19)

Several other global databases have also been used for SHS 2.0 in order to compile better, country- and disease-specific mortality or prevalence data. Specifically, the UNAIDS database for ART coverage(20) and the International Agency for Research on Cancer (IARC)(21) for data on cancer patients by years of diagnosis.

### Selection of SHS-associated conditions

The first step in estimating the SHS burden was to identify the health conditions that most commonly cause SHS from the ICD-10 classification list that require palliative care at the end-of-life due to life-threatening conditions or living with a life-limiting condition (SHS 1.0). The global SHS 2.0 database includes 21 conditions, and these are presented in Table 1 with their corresponding ICD-10 codes and GBD codes. [**Table 1:** 21 conditions included in the global SHS database in ICD-10 codes and GBD disease codes] All 21 groups of conditions include decedent categories, considering that at least a proportion of people dying from those conditions suffer from serious health-related suffering. In addition, non-decedent categories of SHS are included for some of the 21 conditions that: may have been cured but from which SHS persists (drug-resistant tuberculosis, some hemorrhagic fevers such as Ebola, some malignancies, some inflammatory diseases of the central nervous system); from which patients recover but that caused SHS (serious injuries, renal failures, preterm birth complications and birth trauma); with survival with chronic severe disability and with SHS symptoms (cerebrovascular disease, leukemia, congenital malformations, injury, birth trauma, human immunodeficiency viruses / acquired immunodeficiency syndrome (HIV/AIDS), some musculoskeletal disorders, liver diseases); and, have a slowly progressive course (malignancies, dementia, Parkinson’s disease, multiple sclerosis, type-1 diabetes, thalassemia, and sickle cell disorders).

In the original Lancet Commission report, the non-decedents category for 11 conditions were considered. In SHS 2.0, non-decedents categories for four more conditions were added and differentiating factors were used that are important to estimating suffering patterns. Panel 1 provides a detailed description of how decedents and non-decedents in need of palliative care are estimated for each condition as well as key literature and extra databases used to calculate the decedents and non-decedents with SHS. Conditions are ranked using the alphabetical order of their ICD-10 codes.

**Panel 1:**
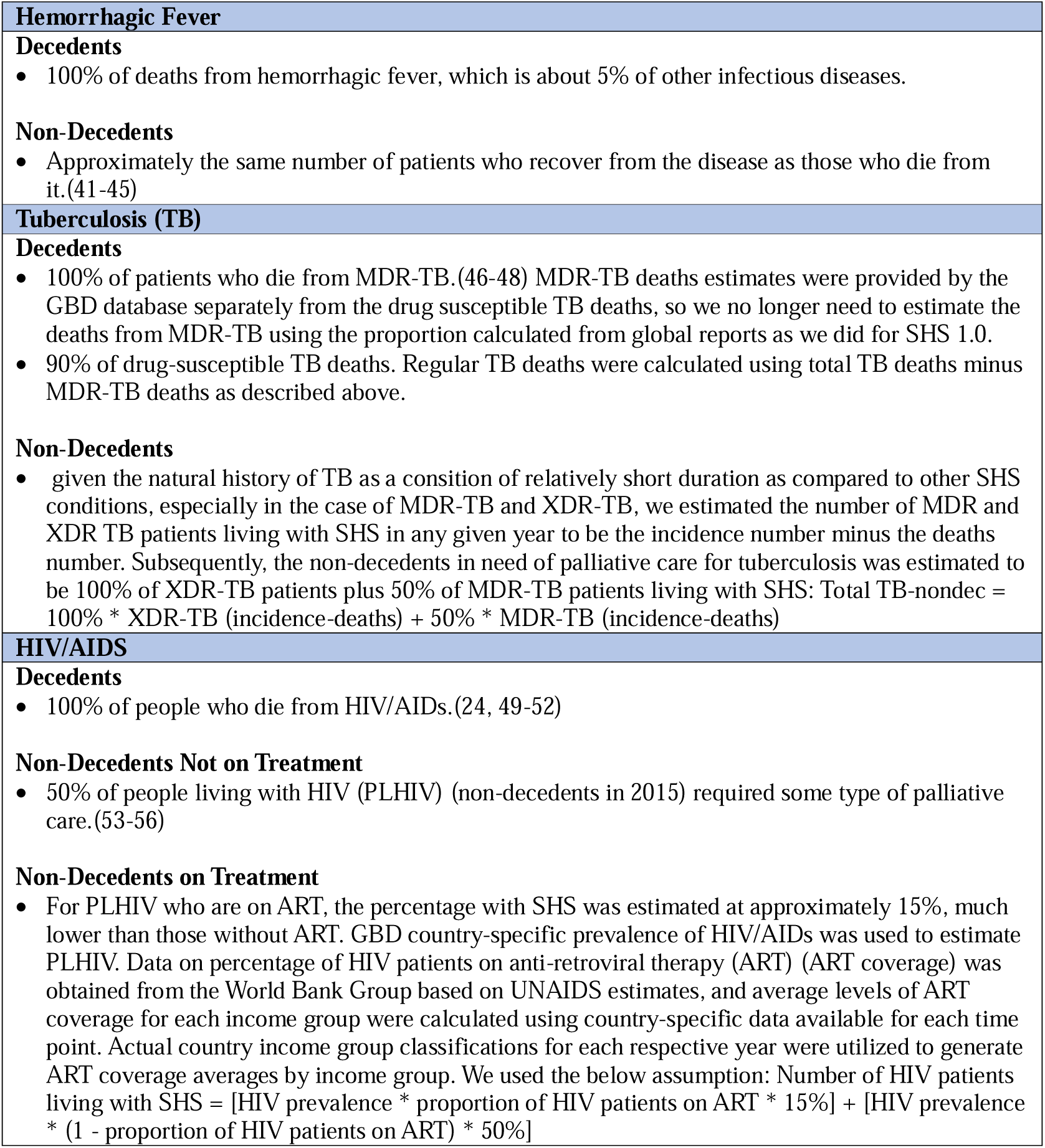
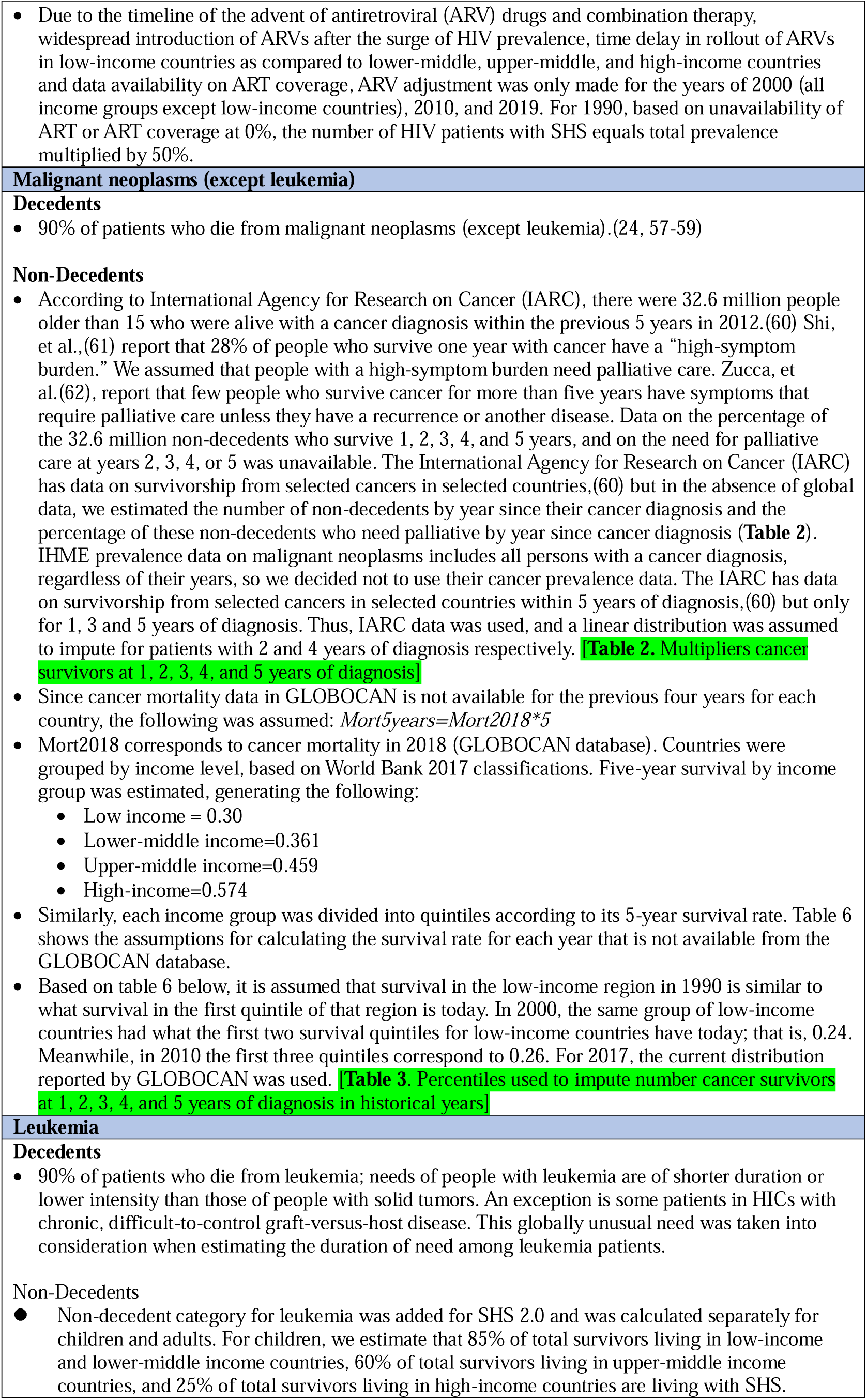
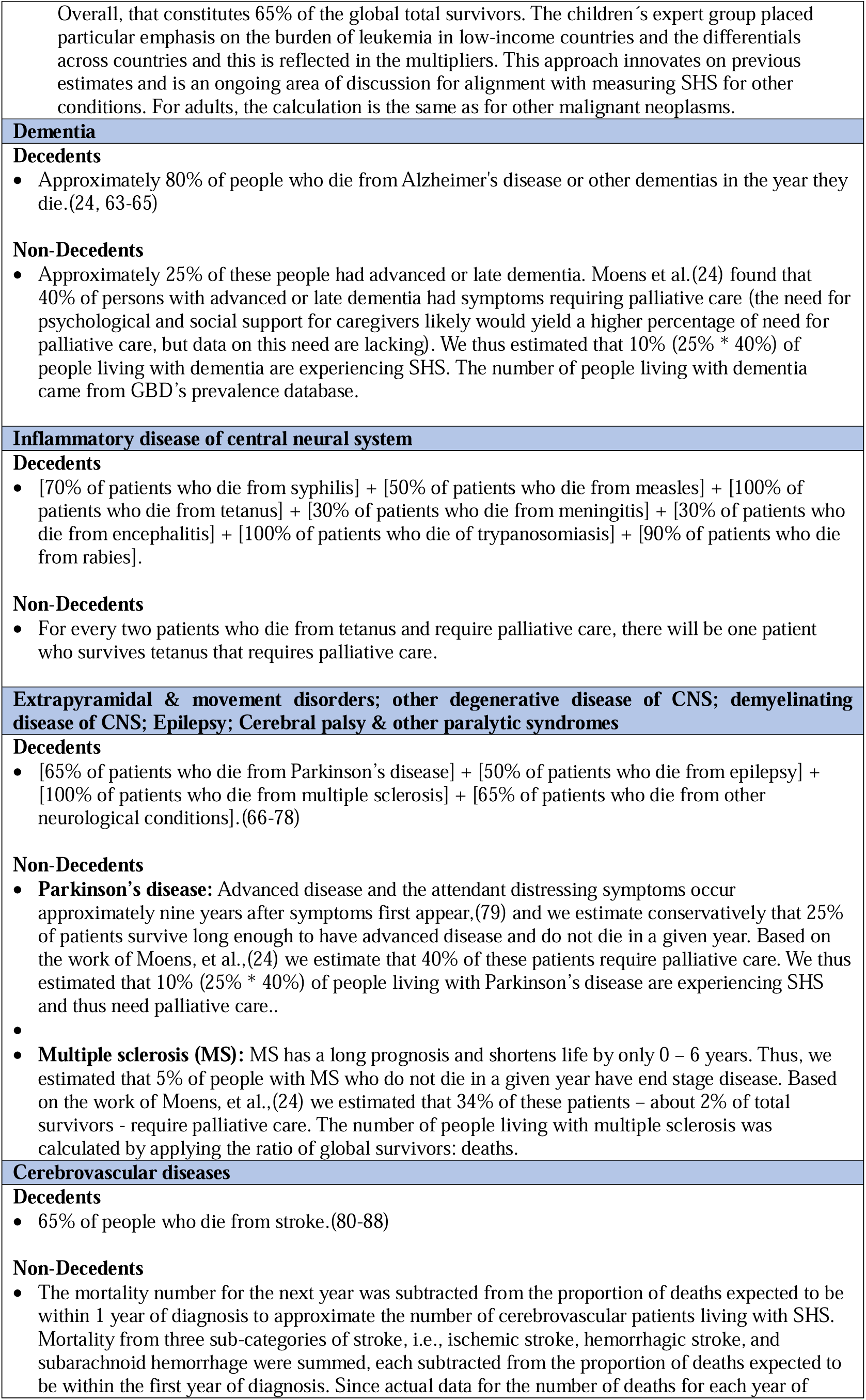
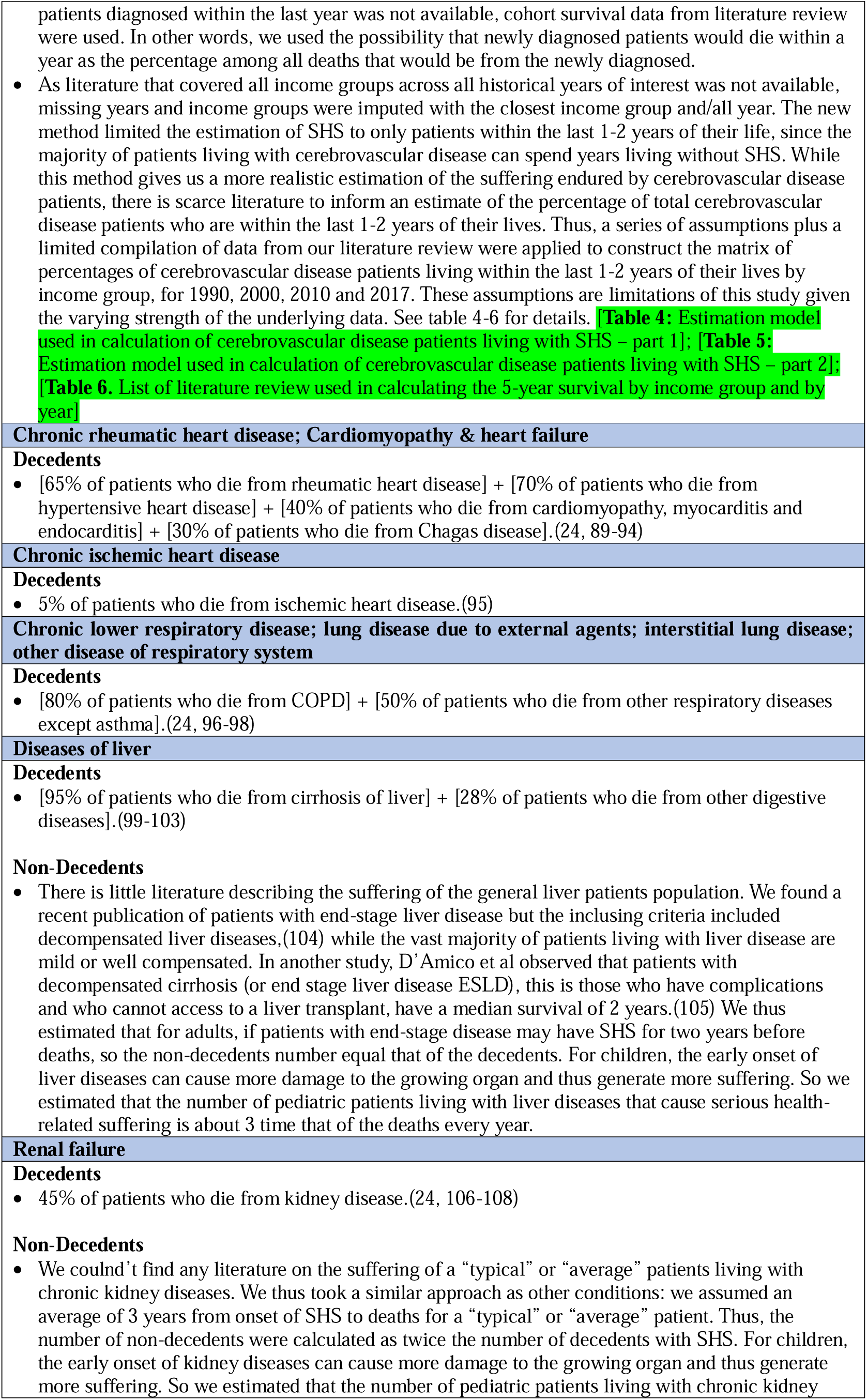
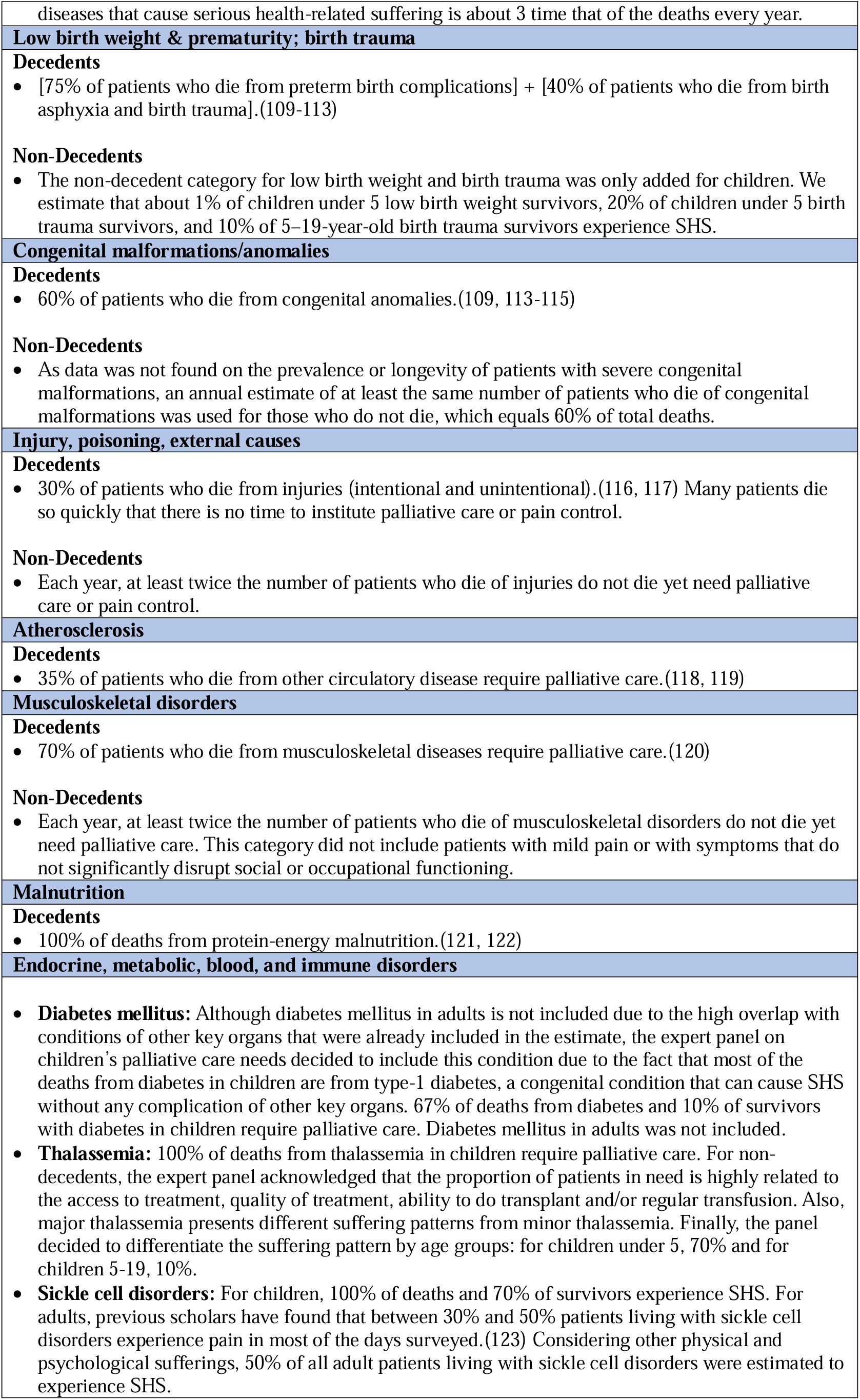
Core assumptions for estimating decedents and non-decedents in need of palliative care.

As the result of the exercise to estimate palliative care needs for children, there was consensus that the following conditions be added due to their substantive contribution to SHS among children for both decedents and non-decedents: 1) diabetes mellitus, 2) sickle cell disorders, 3) thalassemia, and the following conditions for non-decedents categories of: 1) leukemia, 2) liver diseases, 3) chronic kidney diseases, 4) neonatal preterm birth and birth trauma. Hence, while SHS 1.0 included 20 conditions, SHS 2.0 includes 21 groups of conditions with the addition of endocrine, metabolic, blood, and immune disorders which include diabetes mellitus, sickle cell disorders, and thalassemia for decedents and non-decedents.

The review of the case of diabetes in children prompted an overall review of the included conditions. For diabetes mellitus in adults, deaths from sequelae are attributed to the proximal cause and hence considered captured in other conditions included in the SHS database and specifically, cerebrovascular disease, cardiomyopathy and/or heart failure, chronic ischemic heart disease, renal failure, and atherosclerosis. Because deaths from diabetic ketoacidosis or hyperglycemic hyperosmotic non-ketotic syndrome typically result in death so rapidly that there is no time to institute quality palliative care services, these conditions are not included. In the pediatric population, diabetes mellitus is added due to the concerns over pain and suffering caused by type-1 diabetes even in the absence of organ complications.

Efforts to alleviate SHS experienced by a newborn, the assurance of the newborn’s comfort and that of distraught parents should accompany aggressive life-sustaining treatments if they are to reasonably provide more benefit than burden. Palliative care must also be available as an alternative to potentially harmful life-sustaining interventions when a newborn is moribund. Hence, in both SHS 1.0 and SHS 2.0, extremely premature and very low birth weight newborns whose survival is unlikely, and babies born with severe hypoxic ischemic encephalopathy or congenital anomalies not compatible with life are included in the list of SHS conditions.

In both SHS 1.0 and SHS 2.0, leukemia is considered a separate condition than the rest of the malignancies due to its distinctive patients’ demographics and suffering patterns.

### Selection of types or symptoms of suffering

Patients’ suffering varies by type, severity, and duration and a clinically, economically, and strategically useful measure of SHS requires estimation of not only the number of patients who suffer, but also the type of suffering and duration of suffering. Therefore, overarching categories of suffering were identified in SHS1.0 and then within those categories, the types or symptoms were associated with each condition.

Palliative care literature typically divides suffering into four categories – physical, psychological, social, and spiritual – to encompass the full spectrum of human suffering. While the Lancet Commission accepted and adopted all four categories as SHS, the focus was on estimating the prevalence and duration of only physical and psychological categories of suffering and corresponding symptoms. The empirical evidence from published literature or expertise to produce reasonable estimates of the prevalence and duration of each type of social and spiritual suffering were not sufficient.

To estimate SHS as precisely as possible, the Commissiońs expert group identified the most common symptoms of physical and psychological suffering, and then estimated the prevalence and duration of each type of suffering associated with each condition or its treatment. Through literature review and evidence-informed expert consensus building exercises, physical and psychological types of suffering (symptoms), their frequencies and durations for each condition were identified as part of Commission work. See **[Figure 1]** for details. Specifically, the types of physical suffering include: moderate or severe pain, mild pain, weakness, fatigue, shortness of breath, nausea and vomiting, constipation, diarrhea, dry mouth, itching, wounds and bleeding. The types of psychological suffering identified include: – anxiety and worry, depressed mood, delirium or confusion, and dementia with disorientation, agitation, or memory loss. Table 2 summarizes the duration of each type of physical and psychological suffering and appendix table 3 lists the results from the literature review on prevalence of the most commonly reported type of physical suffering among patients with serious, complex, or life0limiting health problem. **[Table 2:** The estimates of duration of each type of physical and psychological suffering by condition]

Most published data on symptom prevalence comes from high or upper-middle income countries where both disease-modifying and palliative treatments are most accessible. Furthermore, most of the literature either focused on physical and psychological symptoms among a single group of patients (such as cancer), or a single symptom (such as pain) in patients with various conditions. Data, mostly from high income countries, indicates that well over 50% of people who die of or live with malignant neoplasms and AIDS experience pain, and that pain is also common among those who live with heart disease, chronic obstructive pulmonary disease (COPD), renal failure, neurologic disease and dementia.(22, 23) Dyspnea (shortness of breath) is especially common among people who live with COPD and heart failure and only slightly less common among those who live with malignant neoplasms and AIDS.(24) Depressed mood and anxiety are widespread among patients with a variety of advanced life-threatening illnesses including metastatic cancer and trauma.(25, 26) There are fewer studies among patients with most other serious, complex, or life-limiting health problems.

Of note, dementia appears both in the list of conditions (Alzheimer’s disease and other primary dementias) and as a symptom of other conditions (HIV/AIDS, cerebrovascular disease, and other neurologic conditions). The term dementia is therefore used in two ways, and the distinction in use of each instance is required.

### Identifying multipliers for each condition

The next step in measuring SHS was to determine the proportion of people with each condition who experience SHS. These are called multipliers. Multipliers are mathematical factors that estimate number of people dying or living with SHS based on different data sources. They reflect different strategies applied in the estimation and are provided separately for decedents and non-decedents. For decedents, the multipliers are always a percentage between 0 and 100%, to be applied to total deaths. For non-decedents, the multipliers take one of the three different forms: 1) a percentage between 0 and 100% to be applied to total number of patients living with the disease; 2) a ratio that can go over 100% to be applied to total deaths; or 3) a ratio that can go over 100% to be applied to total decedents in need of palliative care. See table 4 with more details.

To identify the proportion of people with each condition who experience SHS for the different conditions and sub-conditions and therefore identify appropriate multiplies to use for each, an extensive literature review was conducted for both decedents and non-decedents. Empirical evidence of symptom burden for some conditions was identified, but most studies were conducted in high-income settings. Evidence identified from the literature could not directly be used as multipliers since much of it was focused on patients in a certain stage of care whilst the SHS calculation requires multipliers for both people who die within that year – decedents- and another for people who live with a condition – non-decedents. As a result, empirical evidence on percentage of patients with each condition experiencing SHS from the literature review were summarized and presented as the basis of discussion in various expert consensus building exercises. When estimating the SHS burden of non-decedents, experts were asked to consider the SHS burden of an “average” patient for each condition among all patients living with that condition who are not in their last year of life.

Because SHS 2.0 incorporates analysis across a number of years, it was possible to implement improvements to the multipliers for HIV and tuberculosis (TB). SHS stemming from HIV among non-decedents was differentiated between individuals undergoing anti-retroviral treatment (ART) from those who are not, reflecting how the advent of ART and increased access to such treatment revolutionized care for PLWHIV and in turn, SHS associated with HIV. Furthermore, extensively drug-resistant TB (XDR-TB) is differentiated from multidrug-resistant TB (MDR-TB), because antimicrobial resistance and the rise of XDR TB pose major challenges to treatment of tuberculosis which is different from MDR-TB.

For cancer, SHS2.0 also incorporates data across additional years for the estimation of multipliers. In SHS2.0, unlike for the Commission report, five-year survival data were used to estimate non-decedent SHS for malignant neoplasms and leukemia. The GBD data reports only overall survival and does not further disaggregate by years since diagnosis. Hence, the GBD data were adjusted based on the prevalence and mortality data extracted from the Global Cancer Observatory (GLOBOCAN) 2018 (See Panel 1 and Table 3) that report cancer survivorship for 1,3 and 5 years from diagnosis. [**Table 3**. Multipliers of cancer survivors at 1, 2, 3, 4, and 5 years of diagnosis] A country-specific linearly interpolated trend was applied to estimate prevalence for year 2 and 4 post diagnosis. The approximation of survival was estimated as the ratio between the total deaths and the prevalence in the same period. Lastly, to estimate non-decedent burden for 1990, 2000 and 2010 given that information on 5-year prevalence and survival is not available by year since diagnosis, the GLOBOCAN 2018 data are adjusted using country-income specific quintile distribution data on percentages of all cancer survivors being with each year of diagnosis. (see Table 4 for detail). [**Table 4**. Percentiles used to impute number cancer survivors at 1, 2, 3, 4, and 5 years of diagnosis in historical years]

Cerebrovascular diseases constitute a major component of overall SHS, yet its non-decedent category was a limitation in SHS1.0. For SHS2.0, non-decedent SHS was calculated for patients living in the year prior to their last year of life, assuming that most patients who live for extended periods with this condition do not experience SHS (as the condition is largely asymptomatic until it becomes serious enough to result in death). Still, data are scarce on the proportion of cerebrovascular disease patients in the final years of life and hence with SHS. An estimate of the proportion of patients who are diagnosed and die in the same year was developed based on a literature search focusing on differences by country income level and this was applied to two years of cerebrovascular disease mortality. (see Tables 5, 6 and appendix table 4). [**Table 5**: Estimation model used in calculation of cerebrovascular disease patients living with SHS – part 1] [**Table 6:** Estimation model used in calculation of cerebrovascular disease patients living with SHS – part 2] Because data were not available on the number of deaths per year of patients diagnosed in the last year, a literature search was carried out on the survival of these patients in countries by income level. In other words, the calculation factored in the percentage of newly diagnosed patients that would die within one year as the percentage among all deaths that would occur due to newly diagnosed patients. As literature covering all income groups was not available in all years of interest, i.e. 1990, 2000, 2010 and 2019, missing years and income groups were imputed to the nearest income group and/or to all the year (Tables 8 and 9). The new method limited the estimation of SHS only to patients within the last 1-2 years of their life, since most patients living with cerebrovascular disease can spend years living without SHS. While this method gives us a more realistic estimate of the suffering endured by cerebrovascular disease patients, there is little literature to report an estimate of the percentage of total cerebrovascular disease patients who are in the last 1-2 years of their life (Table 10). Therefore, we applied a series of assumptions plus a limited compilation of data from our literature review to construct the matrix of percentages of cerebrovascular disease patients living within the last 1-2 years of their life by income group, to 1990, 2000, 2010, and 2019. These assumptions are limitations of this study, given the varying strength of the underlying data.

Table 7 presents the multipliers used to calculate SHS for all 21 conditions, separating decedents and non-decedents. [**Table 7**. Multipliers used to calculate SHS burden for 21 conditions]

## III. Data limitations and Future Iterations

The measurement of the global burden of SHS presented in the Lancet Commission report set a precedent and the update to SHS2.0 is an important move forward in measuring the number of people in need of palliative care. However, there are important limitations and there remains work to refine the estimation strategy and hence the estimates.

### Data limitations

First, although a literature review was conducted by condition and symptoms, due to a dearth of reliable empirical data on the types, prevalence, and duration of suffering caused by each SHS associated health condition, both SHS1.0 and 2.0 rely heavily on expert opinion. Moreover, research on palliative care has so far concentrated on Europe and the United States accounting for over 90% of all publications on palliative care but only 15% of the global population. The fact that 85% of the global population produced only 6.5% research publications points to the glaring lack of information on the elements of suffering for the majority of people in the world.(27)

Further, the expert groups are relatively small reflecting limitations in available funding to develop the field of SHS. This makes it especially difficult to develop either disease, region, or country income-specific estimates. The reliance on identifying an “average” patient limits the possibility of exploring regional, cultural or other differences, as well as the effect of providing differential levels of palliative care. The next step in the SHS work is to undertake disease-specific expert panels to refine estimates of people with SHS and especially symptoms and symptom days. This is the focus of research planned for SHS 3.0 and has been piloted for breast cancer and will soon commence on HIV.

Second, there are conditions which generate SHS but are not included in the analysis to-date due to limited scope. For example, chronic paranoid schizophrenia and other severe chronic psychiatric disorders generate severe suffering but are not included in the methods presented here. Another important example is people living in the context of humanitarian crisis,(28) including armed conflict(29) but also climate emergencies, communicable disease outbreaks or those under threat of political, sexual, or ethnic violence who suffer from various types of physical and psychological suffering.

Similarly, our work to date extends to 2019. Estimating the shorter-term SHS that was associated with the coronavirus disease 2019 (COVID-19) pandemic, and the longer-term sequelae for those who suffered the disease should be a key next step in the analysis. This should include the suffering associated with bereavement and the lack of access to palliative care support for caregivers, family members and the community during COVID-19 lockdowns. The wealth of data and publications on the pandemic will make this analysis more feasible.

Family caregivers who experience various kinds of physical, psychological, social, and spiritual suffering as a result of their care work are not included in the estimates. While methods to estimate the types, prevalence, or duration of physical, psychological, social, or spiritual suffering of the main family or informal caregiver have not been within the scope of SHS calculations to-date, this is an important area of future SHS methodological development. Family caregivers typically provide many hours of daily care to patients with serious, chronic, complex, or life-limiting health problems and in many health care settings, especially in LMICs, where they must remain with the patient when admitted to the hospital. Across the world, caregiving work at home and in the communities is predominantly provided by women, and often uncompensated or undercompensated.(30) It has been shown that caregiving can itself represent a source of suffering.(31, 32) Family caregivers may have their own need for palliative care and support in managing bereavement.

Expert opinion provides important information, but a patient-centered approach needs to be included in future work on SHS. Confirmatory research on symptom prevalence and severity with patient- and caregiver-reported real-life data must complement future work. This limitation applies to the symptoms as well as many dimensions of suffering that are important for patients, caregivers, and practitioners. The expert panel identified 11 physical and 4 psychological symptoms, but this is far from an exhaustive list of all possible physical and psychological symptoms patients can experience. Social or spiritual suffering is also not estimated despite being a source of grave concern due to the impact on overall quality of life.(33, 34) In the context of paucity of resources, of poorly organized healthcare systems and of marginalization of large chunks of the population, the impact on the burden of suffering is likely to be considerable.

Further, the quantity of suffering is estimated only in terms of number of people who died from or lived with SHS (SHS 1.0 and SHS 2.0), or the number of symptom days they each experience (SHS 1.0). This approach neglects the intensity or tolerability of suffering experienced. In SHS 3.0, opportunities for understanding the scope and intensity of social and spiritual suffering for patients in need of palliative care will be explored. Gathering patient- and caregiver reported data is the optimal solution to fill in these gaps and should be a priority for donors and foundations interested in improving access to palliative care and achieving the Sustainable Development Goals (SDGs). To date, only a few pilot and exploratory surveys have been undertaken.(35, 36)

Another important area for future work is to determine to what extent suffering can be alleviated with existing practices and techniques at various resource-levels. This also means that the multipliers – percentage of deaths or survivors in need of palliative care by condition – are time-period specific and should change over time based on previously noted endogenous variables, including the change in disease trajectories and their suffering patterns, as health care technologies and systems evolve.

Last but not least, our work looks at one side of the issue: the demand side. It is equally important, if not more, to measure how much of the need for palliative care is fulfilled, by whom, in what quality, and where. Combined with analysis of the actual provision of palliative care, we will be able to identify gaps and provide more tailored policy recommendations.

### Future Iterations

The methods described in this paper are pioneering in the field. However, our exploration has only expanded our vision of the bigger, unknown world, leaving more gaps to be filled with future research. Even the more detailed estimate of “symptom-days” – as opposed to number of people – has limitations as a measure of the burden of SHS experienced by patients in the absence of a method to weigh the tolerability or intensity of each symptom. Specifically, the number of days is calculated for each symptom using the available information on symptom prevalences and duration for each condition. Simple aggregation of days with each symptom may lead to overestimation from double counting, as many patients with advanced disease will suffer from more than one symptom at the same time. As such, the Commission report presented two aggregate indicators to evaluate the total symptom burden: 1) the “at least” SHS-days, which equals the symptom-days from the single most prevalent symptom, in most case, pain, of each condition, and 2) the total symptom days should is the sum total of all symptoms. The actual days of suffering experienced by people with SHS should be a number between these two bounds. Ongoing refinement of the calculation of the number of days of SHS experienced by the population in a given year is a core area for SHS 3.0. Moreover, and as described, it is important to note that the calculation of the number of days of SHS is derived from the calculation of the number of people with SHS, not the other way around. As a contribution to measurement of burden, several “summary indicators” or ways to characterize the suffering experienced by patients were developed. Panel 2 presents these secondary indicators that were constructed for the Lancet Commission report. Another dimension that has not been measured to date is to match SHS to an estimate of palliative care need assessment such as the estimated number of required “palliative care visit-days” – the number of days in which a palliative care provider should see the patient, family or caregiver. Symptom days measures only the days during which the symptom(s) persist(s), regardless of whether a visit by or with a palliative care provider is needed. Severe, refractory, or poorly tolerated symptoms may require daily visits while well-controlled symptoms may require a visit only every 2 to 4 weeks. Indeed, provision of effective palliative care can, and should, reduce the number of symptom days as well as the severity of the symptoms. In doing this, palliative care reduces the SHS burden. This remains an area for future discussion and analysis.

**Panel 2:** Indicator-specific descriptions, assumptions, and limitations.

|  |
| --- |
| **Indicator 1: Total symptom-days by condition** |
| • **Description:** The sum of the symptom-days from each symptom by condition.<br>• **Assumptions and limitations:** No weighting of tolerability of symptoms. Assumption that coinciding symptoms make the suffering worse and thus that the symptom-days from each coinciding symptom should be added together. This assumption generates an overestimation of the total number of days of a patient's suffering. |
| **Indicator 2: AT LEAST symptom-days by condition** |
| • **Description:** The symptom-days of the one symptom of longest duration. This would be the LEAST or minimal number of symptom-days experienced by the patient.<br>• **Assumption and limitation:** Assumes that any other symptoms began and ended during period of the symptom of longest duration. In most cases, this will be an underestimate of the total number of days of a patient's suffering. |
| **Indicator 3: AT LEAST non-pain symptom-days by condition** |
| • **Description:** The symptom-days of the one non-pain symptom of longest duration. This would be the LEAST or minimal number of non-pain symptom-days experienced by the patient.<br>• **Assumption and limitation:** Assumes that any other non-pain symptoms began and ended during period of the non-pain symptom of longest duration. In many cases, this will be an underestimate of the total number of days of a patient's suffering from non-pain symptoms. |
| **Indicator 4: Total pain-days by condition** |
| • **Description:** The sum of mild pain-days and moderate to severe pain-days.<br>• **Assumption and limitation:** The mild pain days do not overlap the moderate to severe pain-days. Thus, this indicator shows total days in pain. However, it does not include any other symptoms. |
| **Indicator 5: Pain plus At LEAST non-pain symptom-days by condition** |
| • **Description:** This indicator was generated by adding the total pain-days and the AT LEAST non-pain symptom-days (indicator 3).<br>• **Assumption and limitation:** This is one possible indicator of the burden of suffering for a patient. |
| **Indicator 6: Total days in need of palliative care by condition** |
| • **Description:** An estimation of days requiring palliative care by condition by palliative care experts with experience treating patients in LMICs using a Delphi process.<br>• **Assumption and limitation:** Based only on the opinion of clinical palliative care experts from LMICs in each region. |

## IV. Discussion

This paper is designed to serve as a reference document for calculating SHS. Detailing the methodology is also intended to promote transparency in ongoing efforts to measure the burden of SHS and to promote wider discourse on the assessment of SHS burden that will inform future iterations of SHS measurement and data strengthening. Improving the science of the measurement of SHS will support policies that increase palliative care access and infrastructure as a component of UHC and improve population health.

The estimates generated from this methodology can be used independently or can serve as an input to the development of composite metrics that compare interventions in terms of suffering averted. Researchers can apply the methods presented using country-specific data (i.e. not GBD estimates, which are used here) to generate national and sub-national calculations of SHS.(37, 38) Researchers can also use our methods to project trends and examine the future scale of the burden of SHS overall or by condition.(39) The SHS burden data is also a necessary input to calculating the cost of an essential package of palliative care services, as introduced by the Lancet Commission.(37)

Data on SHS burden is critical to evaluating health status and as such, for the monitoring and evaluation of health systems performance to achieving universal access to palliative care.(40) The number of people with SHS (calculated without a threshold or cutoff in terms of days of SHS experienced) provides a specific insight on palliative care need – an estimated number of patients that need access to palliative care services. Policymakers and practitioners can be guided by the magnitude of SHS within their countries, the distribution of SHS across conditions, age ranges, and geographical locations, and the corresponding need for palliative care, so that they may examine it against the availability of palliative care service. SHS data are hence useful in assessing the need and efficacy of approaches to health system strengthening and UHC, health reforms or across health insurance schemes. Further, the evidence on need can further the argument for adoption of the packages of palliative care services, as was begun with work on the essential package by the Lancet Commission with the Disease Control Priorities (DCP)-3.(11) The number of days of SHS is therefore also essential and particularly to measure how need must translate into a health system response such as through an essential package of palliative care services.

Acknowledging this and the previously presented limitations, this paper provides a starting point for further scientific inquiry and consensus-building. The methods described in this paper pave the way forward for future research that examines both the demand side—suffering patterns—and the supply side—ways to address them—for people worldwide. With the methodology to measure SHS, as established by this paper, what’s needed next are better tools to measure the responses to relief, building on existing efforts such as DOME. The next step and complement to this paper is another on DOME that begins to identify access to one fact of palliative care – pain relief medicine, plus a paper looking specifically at SHS in children. Matching DOME and SHS provides an indicator of health system performance and progress over time in delivering palliative care and reducing the unmet burden of SHS.

Estimating the burden of SHS should be a continual endeavor to incorporate scientific, societal, economic, and health care system change into the quest to reduce suffering and improve population health. This must include monitoring advances, but also the challenges that pose a risk to human health and quality of life, including climate change, war, and humanitarian crises. The measurement of serious health-related suffering can serve as a basis for promoting people-centered health systems and analyzing progress toward SDG3 and for future iterations of global health goals and the quest for UHC. It also has the potential to change the focus of today’s healthcare system from diseases alone to suffering. The tools shared in this paper and its contributions toward better conceptualization and measurement of the burden and alleviation of SHS should catalyze this work.

## V. Acronyms

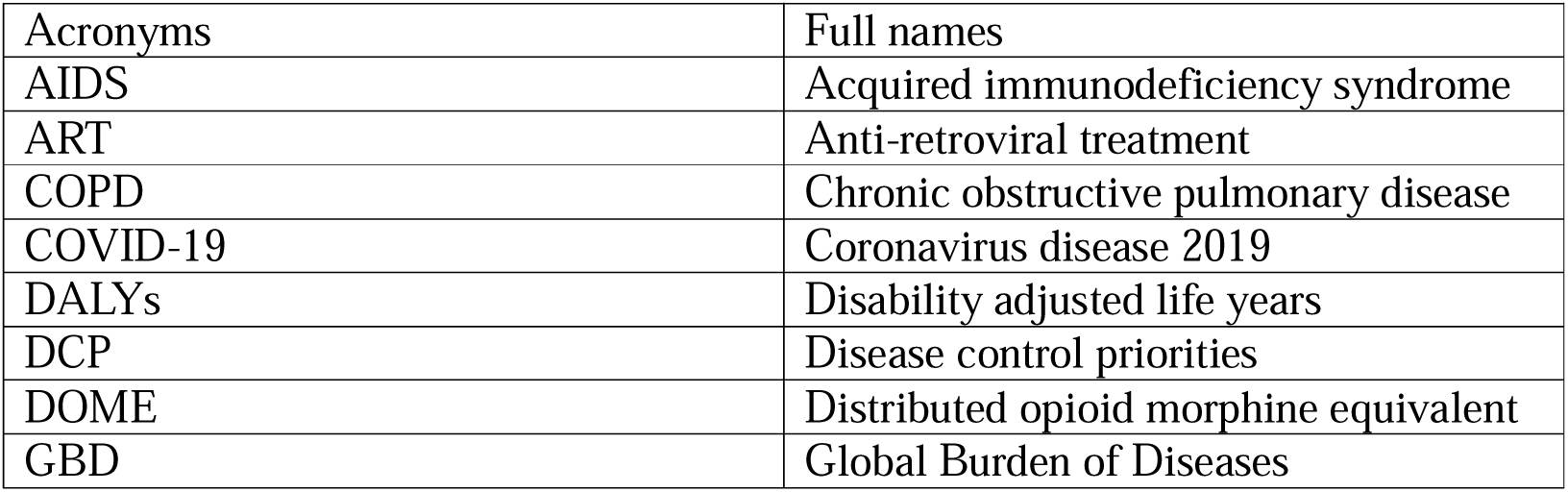
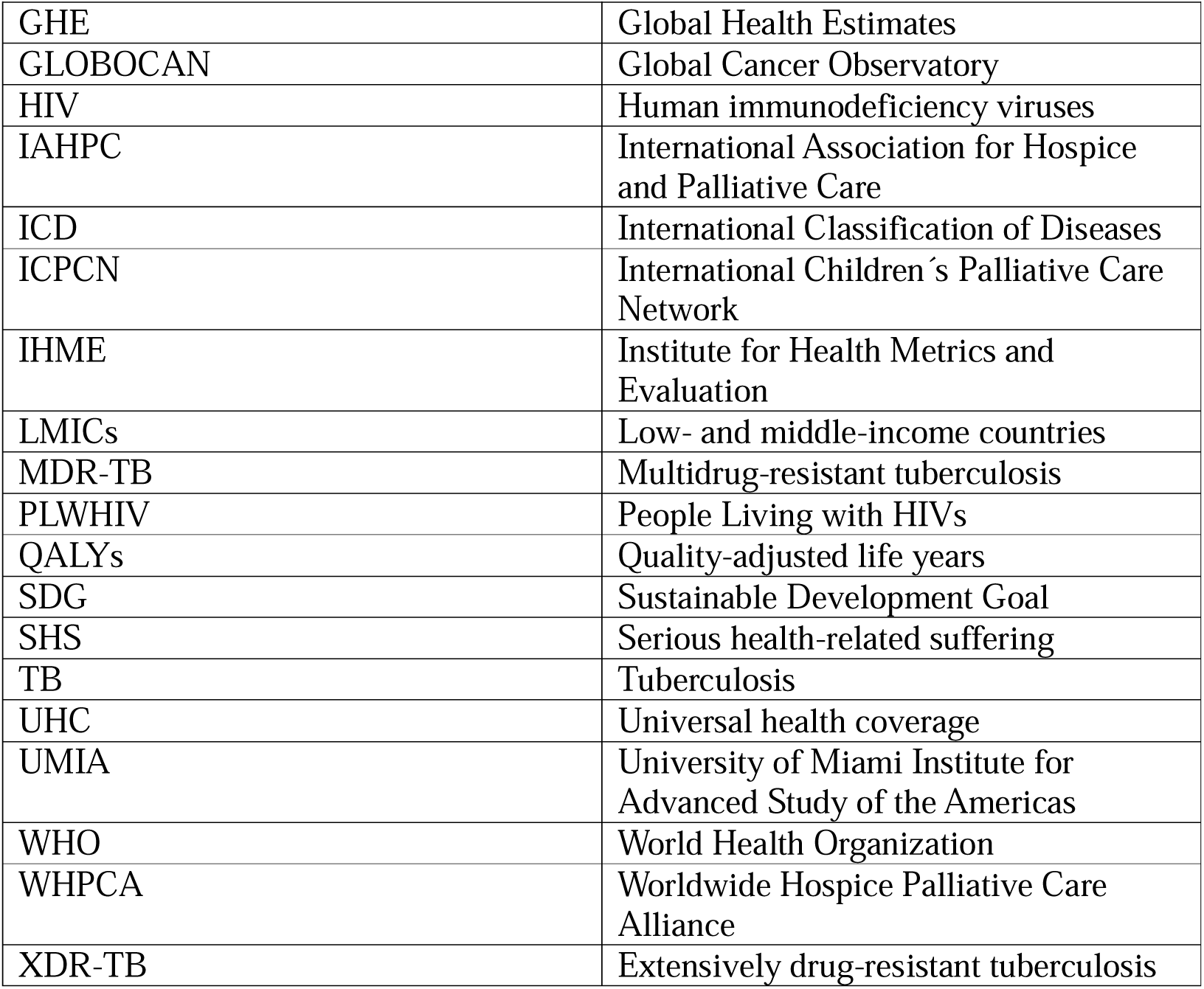

## Data Availability

As this paper is primarily methodological in nature, no additional data were generated for it. However, detailed descriptions of the methodology employed are provided to ensure reproducibility and transparency.

## Acknowledgments

The authors are grateful to the Lancet Commission on Palliative Care and Pain Relief Study Group and acknowledged contributors in the Lancet Commission report for their inputs to an earlier iteration of this work. We would also like to thank all palliative care specialists who contributed to experts panels and related Delphi processes for the Lancet Commission and subsequent pediatric specific expert reviews for SHS 2.0. We thank Kathy Foley for her various inputs to the Lancet Commission and beyond to help make this work a reality. Finally, we thank all individuals who have supported this work in different ways and at varying points.

