## Appendix for "Global Assessment of Palliative Care Need: Serious Health-Related Suffering Measurement Methodology"

**Data Appendix**

**Sectio 1. Additional materials supporting the construction of the Global SHS database:**

| Appendix Table 1. Experts’ consensus building practices undertaken by the LC | | | | |
| --- | --- | --- | --- | --- |
| Dates | Activity | Participants | Location | Content Consulted |
| Between 2015 and 2016 | Experts consultation using in-person one-on-one in-depth interviews | 10 palliative care practitioners from 7 LMICs | Rwanda, Vietnam, Mexico, India, Malaysia, Jamaica, Brazil | Multipliers of SHS, types of symptoms, design of the essential package |
| Aug-16 | Group discussion at a Lancet GAPCPR commission meeting | Commissioners, Scientific Advisory Committee Members, Collaborators | Cuernavaca, Mexico | Multipliers of SHS, frequency and duration of symptoms, design of the essential and augmented package, costing framework, etc. |
| ? | A two-stage Delphi | 18 palliative care practitioners from LMICs | virtually | Design of the essential and the augmented package and days in need of palliative care |
| Jun-21 | Expert panel discussions in a virtual meeting and following up in emails | 8 pediatric palliative care specialists from both HICs and LMICs | virtually | Multipliers of SHS for children, additional conditions for children |

| **Appendix Table2-1. Delphi Process Results from Round1** | | | | | | | | |
| --- | --- | --- | --- | --- | --- | --- | --- | --- |
| **Participant Number** | **1** | **2** | **3** | **4** | **5** | **6** | **7** | **8** |
| **1. Should any medicines, equipment, or social supports be added to, or removed from, the essential/highest priority package? If yes, please provide details below.** | | | | | | | | |
| **Please provide information on what medicines should be added to or removed from the essential/highest priority package:** | should add codein, tramadol, carbamazepine | added: Diclofenac tabs,Tramadol tabs | It would be important to add Acid Tranexamic in my opinion, I do not see so important Fluoxetin or other SSRI if we have Amitriptillin in the list. | added: Midazolan I.V. and Lorazepam oral, Ketorolaco I.V., Metamizol I.V. Olanzapina IV and oral. Paracetmol parenteral. Removed: Diazepam | Nebulization medicines for breathlessness | Should be added: Fentanyl patch and Octeotride parenteral / Should be removed: Loperamide | 1. Entacyd (with Simethicone) oral 2. Body Lotion 3. Antifungal oral 4. Anti fungal ointment |  |
| **Please provide information on what equipment should be added to or removed from the essential/highest priority package:** | convert a suspention cot | NonSterile Gloves, thermometer, BP machine, stethoscope | Antidecubitus materials or medications like Duoderma and Colostoma I do see as high priority to be added | equipment of paracentesis,equipment healing of ulcers and wounds butterfly needles for subcutaneously | nebulizer | Should be added: wheelchairs and 3 positions beds. Bathroom and bath chairs. / Should be removed: Nasogastric drainage or feedin tube | Small kit containing few gauze pads, 1 nail cutter, 1 scissor, 3 syringes - 3 cc, 5 cc, 10 cc, one adhesive micropore |  |
| **Please provide information on what social support should be added to or removed from the essential/highest priority package:** | enough | None | I don't have any suggestion in this respect | chapel, hostel for families, relaxation room for families, | Ok | Costa Rica provides a special license (Law 7756) for the patient relative who takes care of the terminal patient, during the time he stays alive at home / Should be removed: Cash payment and housing | Education support for 1/2 dependants of the patient |  |
| **2. Should any medicines, equipment, or social supports be added to, or removed from, the augmented (second tier) package? If yes, please provide details below.** | | | | | | | | |
| **Please provide information on what medicines should be added to or removed from the augmented package:** | should add endocrine therapy | added: fentanyl patches, Imodium tabs, Multivitamin, Ion tabs | Not anything to say | added: Midazolan I.V. and Lorazepam oral, Paracetmol parenteral. Removed: Diazepam | Some alternative medicines like prednisolone,pheniramine,spironolactone, | Should be added: Fentanyl patch and Octeotride parenteral / Should be removed: Loperamide | Same as 1 |  |
| **Please provide information on what equipment should be added to or removed from the augmented package:** | convert a suspention cot | added: Wounds dressing packages, NonSterile Gloves | Not anything to say | equipment of paracentesis,equipment healing of ulcers and wounds butterfly needles for subcutaneously | Pulse oximetry | Should be added: wheelchairs and 3 positions beds. Bathroom and bath chairs. / Should be removed: Nasogastric drainage or feedin tube | Same as 1 |  |
| **Please provide information on what social support should be added to or removed from the augmented package:** | should add psychologic support |  | Not anything to say | chapel, hostel for families, relaxation room for families | Ok | Costa Rica provides a special license (Law 7756) for the patient relative who takes care of the terminal patient, during the time he stays alive at home / Should be removed: Cash payment and housing | Same as 1 |  |
| **3. Please state any suggestions, comments, or concerns you have about the essential/highest priority package.** | delivering and using morphine at home | Very good list especially for LMICs needs in PC | ondansetron oral, scopolamine parenteral | Really it would be great if our governments will adopt this package, it would be great for patients and families also for our teams and for the health system | in my country is very important human resource, often exists but there is no way to pay it. Institutions could be responsible for paying the salaries of doctor, nurse and psychologist in palliative care. We also have a small space in the units to meet. | This is highly appreciating work. But I am 100% sure that this will not be universally accessible by everyone everywhere by 2020. | Suggestion: The creation of a "goverment opiod law deliver" for the whole population of terminal patients in low/medium income countries. | Essential/highest priority package should be distributed through a committee formatted by government and non-government members |
| **4. Please state any suggestions, comments, or concerns you have about the augmented package.** | control breakthrough pain with rapid onset opioids | I would suggest to add package of education program & activities ( counseling, nutrition education, advocacy...) | Radiology tests, CT Scan or MRI would be very important when these are indicated or needed | in my country is very important human resource, often exists but there is no way to pay it. Institutions could be responsible for paying the salaries of doctor, nurse and psychologist in palliative care. We also have a small space in the units to meet. | There should be action oriented plan! | The principal concern is about the use of Nasogastric drenaige (an invasive method in these stage of a patien illnes) as an option for nutrition. | Same as above |  |
| **5. For questions 5.1-5.22, please provide your estimated range of the number of days that patients with each condition noted below would require palliative care for any reason. NO RESEARCH REQUIRED, ONLY YOUR EXPERT OPINION.** | OK but the questions need to be discussed in a session to define at which moment "Palliative care" should start. LMICs & HICs have different approaches. | 15 días | During the whole process of the patients decease |  |  |  |  |  |
| **5.1 Hemorrhagic fevers (includes patients who do not die)** | | | | | | | | |
| **Lower bound for average number of days requiring palliative care** | 3 | 7 | 1 | 7 | 2 | 7 | 15 | 10 |
| **Upper bound for average number of days requiring palliative care** | 7 | 30 | 10 | 60 | 12 | 90 | 60 | 30 |
| **5.2 Tuberculosis (TB) (Death from TB)** | | | | | | | | |
| **Lower bound for average number of days requiring palliative care** | N/A | 15 | 30 | 60 | 15 | 90 | 60 | 180 |
| **Upper bound for average number of days requiring palliative care** | N/A | 90 | 120 | 180-270 | 60 | 365 | 180 | 730 |
| **5.3 Tuberculosis (Death from MDR TB / XDR TB)** | | | | | | | | |
| **Lower bound for average number of days requiring palliative care** | N/A | 10 | 30 | few weeks | 2 | 30 | 240 | 180 |
| **Upper bound for average number of days requiring palliative care** | N/A | 60 | 90 | few months | 12 | 180 | 1825-3650 | 730 |
| **5.4 Tuberculosis (On-treatment for MDR TB / XDR TB (outcome uncertain, both cure and death possible)** | | | | | | | | |
| **Lower bound for average number of days requiring palliative care** | N/A | 8 | 30 | 30 | 2 | 180 | 730 | 180 |
| **Upper bound for average number of days requiring palliative care** | N/A | 90 | 180 | 180 | 12 | 365 | 3650 | 1000 |
| **5.5 HIV/AIDS** | | | | | | | | |
| **Lower bound for average number of days requiring palliative care** | N/A | 15 | 30 | 180 | 2 | 180 | 730 | 1000 |
| **Upper bound for average number of days requiring palliative care** | N/A | 90 | 180 | 365 | 10 | 365 | 3650 | 1000 |
| **5.6 Malignant neoplasms (Death from malignant neoplasms)** | | | | | | | | |
| **Lower bound for average number of days requiring palliative care** | 5 | 8 | 30 | 180 | 5 | 30 | 120 | 10 |
| **Upper bound for average number of days requiring palliative care** | 30 | 30 | 90 | 365 | 20 | 365 | 730 | 100 |
| **5.7 Malignant neoplasms (Survivors of malignant neoplasms)** | | | | | | | | |
| **Lower bound for average number of days requiring palliative care** | 7 | 7 | 30 | 90 | 10 | 365 | 730 | 10 |
| **Upper bound for average number of days requiring palliative care** | 90 | 60 | 180 | 730 | 60 | 1825 | 3650 | 1500 |
| **5.8 Leukemia** | | | | | | | | |
| **Lower bound for average number of days requiring palliative care** | 5 | 8 | 30 | 180 | 15 | 30 | 365 | 10 |
| **Upper bound for average number of days requiring palliative care** | 30 | 60 | 180 | 365 | 120 | 90 | 1825 | 1000 |
| **5.9 Dementia** | | | | | | | | |
| **Lower bound for average number of days requiring palliative care** | 30 | 10 | 30 | 180 | 30 | 30 | 1825 | 30 |
| **Upper bound for average number of days requiring palliative care** | 180 | 60 | 365 | 730-1095 | 90 | 180 | 3650 | 2000 |
| **5.10 Inflammatory disease of CNS** | | | | | | | | |
| **Lower bound for average number of days requiring palliative care** | 14 | 7 | 30 | 7-14 | 90 | 30 | 1825 | 10 |
| **Upper bound for average number of days requiring palliative care** | 30 | 30 | 365 | 150-180 | 180 | 90 | 3650 | 30 |
| **5.11 Following disease types: (a) Extrapyramidal & movement disorders, (b) Other degenerative diseases of the CNS, (c) Demyelinating  disease of the CNS, (d) Epilepsy, (e) Cerebral palsy & other paralytic syndromes** | | | | | | | | |
| **Lower bound for average number of days requiring palliative care** | 20 | 30 | 30 | 30 | 90 | 180 | 1825 | 30 |
| **Upper bound for average number of days requiring palliative care** | 90 | 120 | 365 | 730-1095 | 180 | 365 | 3650 | 1000 |
| **5.12 Cerebrovascular disease** | | | | | | | | |
| **Lower bound for average number of days requiring palliative care** | 20 | 12 | 30 | 7 | 15 | 90 | 1825 | 30 |
| **Upper bound for average number of days requiring palliative care** | 90 | 30 | 180 | 120-150 | 30 | 180 | 3650 | 1000 |
| **5.13 Following disease types: (a) Chronic rheumatic heart diseases, (b) Cardiomyopathy & Heart failure** | | | | | | | | |
| **Lower bound for average number of days requiring palliative care** | 15 | 7 | 1 | 14-21 | 30 | 30 | 365 | 30 |
| **Upper bound for average number of days requiring palliative care** | 60 | 21 | 90 | 365 | 90 | 90 | 3650 | 1000 |
| **5.14 Chronic ischemic heart disease** | | | | | | | | |
| **Lower bound for average number of days requiring palliative care** | 15 | 7 | 30 | 14-21 | 90 | 180 | 365 | 30 |
| **Upper bound for average number of days requiring palliative care** | 30 | 20 | 180 | 365 | 180 | 365 | 3650 | 3000 |
| **5.15 Following disease types: (a) Chronic lower respiratory disease, (b) Lung disease due to external agents, (c) Interstitial lung disease, (d) Other diseases of the respiratory system** | | | | | | | | |
| **Lower bound for average number of days requiring palliative care** | 7 | 30 | 30 | 30 | 15 | 30 | 1095 | 30 |
| **Upper bound for average number of days requiring palliative care** | 60 | 120 | 700 | 365 | 90 | 365 | 3650 | 3000 |
| **5.16 Diseases of the liver** | | | | | | | | |
| **Lower bound for average number of days requiring palliative care** | 7 | 30 | 30 | 30 | 15 | 180 | 730 | 30 |
| **Upper bound for average number of days requiring palliative care** | 60 | 120 | 120 | 270 | 60 | 365 | 3650 | 3000 |
| **5.17 Renal failure** | | | | | | | | |
| **Lower bound for average number of days requiring palliative care** | 7 | 30 | 30 | 30 | 7 | 30 | 730 | 30 |
| **Upper bound for average number of days requiring palliative care** | 90 | 120 | 90 | 180 | 15 | 180 | 1825 | 3000 |
| **5.18 Following disease types: (a) Low birth weight & prematurity, (b) Birth trauma** | | | | | | | | |
| **Lower bound for average number of days requiring palliative care** | N/A | 14 | 1 | 30 | no data | 7 | 180 | 30 |
| **Upper bound for average number of days requiring palliative care** | N/A | 30 | 30 | some months | no data | 90 | 365 | 4000 |
| **5.19 Congenital malformations** | | | | | | | | |
| **Lower bound for average number of days requiring palliative care** | N/A | 5 | 1 | 30 | no data | 180 | 365 | 30 |
| **Upper bound for average number of days requiring palliative care** | N/A | 14 | 30 | several years | no data | 365 | 7300 | life long |
| **5.20 Injury, poisoning, external causes** | | | | | | | | |
| **Lower bound for average number of days requiring palliative care** | 7 | 7 | 1 | 7 | 15 | 7 | 365 | 10 |
| **Upper bound for average number of days requiring palliative care** | 30 | 20 | 30 | 60-90 | 30 | 30 | 1825 | 30 |
| **5.21 Atherosclerosis** | | | | | | | | |
| **Lower bound for average number of days requiring palliative care** | N/A | 30 | 30 | 30 | 180 | 90 | 365 | 30 |
| **Upper bound for average number of days requiring palliative care** | N/A | 60 | 700 | 365 | 365 | 180 | 3650 | 3000 |
| **5.22 Musculoskeletal disorders** | | | | | | | | |
| **Lower bound for average number of days requiring palliative care** | 15 | 30 | 30 | 30 | 90 | 90 | 1095 | 30 |
| **Upper bound for average number of days requiring palliative care** | 30 | 120 | 700 | several years | 180 | 365 | 3650 | 1000 |

| **Appendix Table2-2. Delphi Process Results from Round2** | | | | |
| --- | --- | --- | --- | --- |
| **DELPHI ROUND 2 RESULTS:** | **Mean** | **SD** | **Mean** | **SD** |
| **DURATION PALLIATIVE CARE IS REQUIRED** | **lower bound (days)** | **+/-** | **upper bound(days)** | **+/-** |
| **CONDITION** |  |  |  |  |
| **Hemorrhagic fevers (includes patients who do not die)** | **13** | **12** | **40** | **35** |
| **Tuberculosis (TB) (Death from TB)** | **80** | **68** | **184** | **133** |
| **Tuberculosis (Death from MDR TB / XDR TB)** | **75** | **31** | **168** | **97** |
| **Tuberculosis (On-treatment for MDR TB / XDR TB (outcome uncertain, both cure and death possible)** | **83** | **39** | **288** | **283** |
| **HIV/AIDS** | **150** | **117** | **316** | **216** |
| **Malignant neoplasms (Death from malignant neoplasms)** | **44** | **26** | **178** | **76** |
| **Malignant neoplasms (Survivors of malignant neoplasms)** | **195** | **140** | **768** | **634** |
| **Leukemia** | **85** | **56** | **249** | **132** |
| **Dementia** | **148** | **79** | **599** | **328** |
| **Inflammatory disease of CNS** | **78** | **32** | **241** | **111** |
| **Following disease types: (a) Extrapyramidal & movement disorders, (b) Other degenerative diseases of the CNS, (c) Demyelinating  disease of the CNS, (d) Epilepsy, (e) Cerebral palsy & other paralytic syndromes** | **173** | **111** | **578** | **361** |
| **Cerebrovascular disease** | **150** | **117** | **419** | **229** |
| **Following disease types: (a) Chronic rheumatic heart diseases, (b) Cardiomyopathy & Heart failure** | **95** | **62** | **433** | **327** |
| **Chronic ischemic heart disease** | **83** | **67** | **349** | **275** |
| **Following disease types: (a) Chronic lower respiratory disease, (b) Lung disease due to external agents, (c) Interstitial lung disease, (d) Other diseases of the respiratory system** | **83** | **67** | **696** | **800** |
| **Diseases of the liver** | **85** | **66** | **433** | **327** |
| **Renal failure** | **88** | **65** | **436** | **219** |
| **Following disease types: (a) Low birth weight & prematurity, (b) Birth trauma** | **32** | **22** | **443** | **705** |
| **Congenital malformations** | **123** | **111** | **430** | **332** |
| **Injury, poisoning, external causes** | **27** | **7** | **202** | **98** |
| **Atherosclerosis** | **88** | **65** | **324** | **83** |
| **Musculoskeletal disorders** | **83** | **67** | **479** | **291** |

| **Appendix Table 3. Literature review on prevalence of the most commonly reported types of physical suffering among patients with serious, complex or life-limiting health problems.** | | | | | | | | | | | | | | | | | | | | | | | |
| --- | --- | --- | --- | --- | --- | --- | --- | --- | --- | --- | --- | --- | --- | --- | --- | --- | --- | --- | --- | --- | --- | --- | --- |
|  | Pain Mild | | | Pain Mod/Severe | | Dyspnea | | Fatigue | | Weakness | | Nausea and/or vomiting | | Diarrhea | | Constipation | | Dry Mouth | | Pruritus | | Bleeding | |
|  | p | ref | | p | ref | p | ref | p | ref | p | ref | p | ref | p | ref | p | ref | p | ref | p | ref | p | ref |
| Hemorrhagic fevers | 81% | Qin, E, 2015 | |  |  | 14.30% | Qin, E, 2015 | 71.40% | Qin, E, 2015 | 100% | Qin, E, 2015 | 24.2% - 57% | Qin, E, 2015; Thomas, E, 2007 | 66.70% | Qin, E, 2015 |  |  |  |  | 27.60% | Thomas, E, 2007 | 8-57% | Qin, E, 2015; Thomas, E, 2007 |
| TB/M/XDR TB |  |  | |  |  | 5.6-30% | Marais, B. J, 2005; Bark, C. M. 2011 | 16.70% | Marais, B. J, 2005 |  |  |  |  |  |  |  |  |  |  |  |  |  |  |
| HIV disease | 30-98% | VietNam National Palliative Care Report; Harding, R., 2012; Solano, J. P., 2006; Moens, K., 2014; Vogl, D., 1999 | | 64-64.70% | VietNam National Palliative Care Report; Harding, R., 2012 | 11-62% | Harding, R., 2012; Solano, J. P., 2006; Vogl, D., 1999 | 43-95% | Harding, R., 2012; Solano, J. P., 2006; Moens, K., 2014 | 71.9-85.5% | Harding, R., 2012; Vogl, D., 1999 | 21-60.7% | VietNam National Palliative Care Report; Harding, R., 2012; Solano, J. P., 2006; Moens, K., 2014; Vogl, D., 1999 | 24.6-90% | Harding, R., 2012; Solano, J. P., 2006; Moens, K., 2014; Vogl, D., 1999 | 34-38.1% | Harding, R., 2012; Solano, J. P., 2006; Vogl, D., 1999 | 61.6-67.6% | Harding, R., 2012; Vogl, D., 1999 | 49-58.9% | VietNam National Palliative Care Report; Harding, R., 2012; Vogl, D., 1999 |  |  |
| Malignant neoplasms (except C91-95) | 30-96% | VietNam National Palliative Care Report; Teunissen, S. C.,2007; Solano, J. P., 2006; Moens, K., 2014; Tranmer, J. E.,2003 | | 66% | VietNam National Palliative Care Report | 10-77% | Teunissen, S. C., 2007; Dudgeon, D. J., 2001; Solano, J. P., 2006; Moens, K., 2014; Tranmer, J. E., 2003 | 23-100% | Teunissen, S. C., 2007; Solano, J. P., 2006; Moens, K., 2014 | 60% | Teunissen, S. C., 2007 | 2-78% | VietNam National Palliative Care Report; Teunissen, S. C., 2007; Solano, J. P., 2006; Moens, K., 2014; Tranmer, J. E., 2003 | 3-29% | Teunissen, S. C., 2007; Solano, J. P., 2006; Moens, K., 2014; Tranmer, J. E., 2003 | 4-65% | Teunissen, S. C., 2007; Solano, J. P., 2006; Moens, K., 2014; Tranmer, J. E., 2003 | 40-82% | Teunissen, S. C., 2007; Tranmer, J. E., 2003 | 0-24% | VietNam National Palliative Care Report; Teunissen, S. C., 2007; Tranmer, J. E., 2003 | 15% | Teunissen, S. C., 2007 |
| Leukemia | 28.2-54.3% | Collins, J. J., 2000; Huijer, H. A. S., 2013 | |  |  | 21.90% | Collins, J. J., 2000 | 15.4-63% | Collins, J. J., 2000; Huijer, H. A. S., 2013 |  |  | 43.8+12.5% - 39.1+23.9% | Collins, J. J., 2000; Huijer, H. A. S., 2013 | 21.90% | Collins, J. J., 2000 | 6.30% | Collins, J. J., 2000 | 21.9-26.1% | Collins, J. J., 2000; Huijer, H. A. S., 2013 | 15.4-40.6% | Collins, J. J., 2000; Huijer, H. A. S., 2013 |  |  |
| Dementia | 14-63% | Moens, K., 2014 | |  |  | 12-52% | Moens, K., 2014 | 22% | Moens, K., 2014 |  |  | 8% | Moens, K., 2014 |  |  | 40% | Moens, K., 2014 |  |  |  |  |  |  |
| Degeneration of CNS | 42-85% | Moens, K., 2014 |  | |  | 26% | Moens, K., 2014 | 42-80% | Moens, K., 2014 | |  | 26% | Moens, K., 2014 |  |  | 24-46% | Moens, K., 2014 | |  |  |  |  |  |
| Non-ischemic heart disease | 14-78% | Solano, J. P., 2006; Moens, K., 2014 | |  |  | 18-88% | Ahmed, A., 2006; Solano, J. P., 2006; Moens, K., 2014 | 42-82% | Solano, J. P., 2006; Moens, K., 2014 |  |  | 2-48% | Solano, J. P., 2006; Moens, K., 2014 | 12% | Solano, J. P., 2006 | 12-42% | Solano, J. P., 2006; Moens, K., 2014 |  |  |  |  |  |  |
| Chronic ischemic heart disease | 41-77% | Solano, J. P., 2006 | | |  | 60-88% | Solano, J. P., 2006 | 69-82% | Solano, J. P., 2006 | |  | 17-48% | Solano, J. P., 2006 | 12% | Solano, J. P., 2006 | 38-42% | Solano, J. P., 2006 | |  |  |  |  |  |
| Lung Diseases | 21-77% | Solano, J. P., 2006; Moens, K., 2014 | |  |  | 56-98% | Solano, J. P., 2006; Moens, K., 2014 | 32-90% | Solano, J. P., 2006; Moens, K., 2014 |  |  | 4% | Moens, K., 2014 |  |  | 12-44% | Solano, J. P., 2006; Moens, K., 2014 |  |  |  |  |  |  |
| N17-19: Renal failure | 11-83% | Solano, J. P., 2006; Murtagh, F. E., 2010; Moens, K., 2014 | |  |  | 11-82% | Solano, J. P., 2006; Murtagh, F. E., 2010; Moens, K., 2014 | 13-100% | Solano, J. P., 2006; Murtagh, F. E., 2010; Moens, K., 2014 |  |  | 8-59% | Solano, J. P., 2006; Murtagh, F. E., 2010; Moens, K., 2014 | 8-36% | Solano, J. P., 2006; Murtagh, F. E., 2010; Moens, K., 2014 | 8-70% | Solano, J. P., 2006; Murtagh, F. E., 2010; Moens, K., 2014 | 69% | Murtagh, F. E., 2010 | 84% | Murtagh, F. E., 2010 |  |  |

| **Appendix Table 4. List of literature review used in calculating the 5-year survival by income group and by year** | | | | | | | | |
| --- | --- | --- | --- | --- | --- | --- | --- | --- |
| **Article** | **Year** | **Population** | **Subtype** | **N** | **Probability of death within 28 days** | | **Probability of death at 1 year** | |
| [Anderson, C. S., Jamrozik, K. D., Broadhurst, R. J., & Stewart-Wynne, E. G. (1994)](https://www.ahajournals.org/doi/pdf/10.1161/01.STR.25.10.1935) | 1994 | Perth, Western Australia | Ischemic | 247 | 30 | 12% | 63 | 26% |
|  |  |  | Heomrragic | 44 | 13 | 30% | 17 | 39% |
|  |  |  | Subcranial | 18 | 6 | 33% | 8 | 44% |
| [Dennis, M. S., Burn, J. P., Sandercock, P. A., Bamford, J. M., Wade, D. T., & Warlow, C. P. (1993)](https://www.ahajournals.org/doi/abs/10.1161/01.str.24.6.796) | 1993 | Comunidad de Oxfordshire, UK | Ischemic | 545 | 57 | 10% | 125 | 23% |
|  |  |  | Heomrragic | 66 | 34 | 52% | 41 | 62% |
|  |  |  | Subcranial | 33 | 15 | 45% | 16 | 48% |
| [Hankey, G. J., Jamrozik, K., Broadhurst, R. J., Forbes, S., Burvill, P. W., Anderson, C. S., & Stewart-Wynne, E. G. (2000)](https://www.ahajournals.org/doi/full/10.1161/01.STR.31.9.2080) | 2000 | Perth, Western Australia |  |  |  | 12% |  |  |
|  |  |  |  |  |  | 32% |  |  |
|  |  |  |  |  |  | 38% |  |  |
|  | 2006 | Bosnia-Hz | Ischemic | 613 | 171 | 28% | 252 | 41% |
|  |  |  | Heomrragic | 223 | 130 | 58% | 138 | 62% |
|  |  |  | Subcranial |  |  |  |  |  |
| [Sacco, R. L., Wolf, P. A., Kannel, W. B., & McNamara, P. M. (1982)](https://www.ahajournals.org/doi/abs/10.1161/01.STR.13.3.290) | 1982 | The Framingham study | Ischemic | 111 | 17 | 15% |  |  |
|  |  |  | Heomrragic | 10 | 8 | 82% |  |  |
|  |  |  | Subcranial | 16 | 7 | 46% |  |  |
| petty, G. W., Brown Jr, R. D., Whisnant, J. P., Sicks, J. D., O’fallon, W. M., & Wiebers, D. O. (2000) | 1996 | Rochester, Minnesota | Ischemic |  |  | 16% |  |  |
|  |  |  | Heomrragic | |  | 58% |  |  |
|  |  |  | Subcranial |  |  | 37% |  |  |
| [Brønnum-Hansen, H., Davidsen, M., & Thorvaldsen, P. (2001)](https://www.ahajournals.org/doi/full/10.1161/hs0901.094253) |  |  | Ischemic | 4162 |  | 28% |  | 41% |
|  |  |  | Heomrragic |  |  |  |  |  |
|  |  |  | Subcranial |  |  |  |  |  |
| Hardie, K., Hankey, G. J., Jamrozik, K., Broadhurst, R. J., & Anderson, C. (2003) | 2003 | Perth, Western Australia | Ischemic | 173 | 16 | 9% | 45 | 26% |
|  |  |  | Heomrragic | 32 | 12 | 38% | 14 | 44% |
|  |  |  | Subcranial | 10 | 5 | 50% | 5 | 50% |
| [Feng, W., Hendry, R. M., & Adams, R. J. (2010)](https://n.neurology.org/content/74/7/588.short) | 2010 | South Carolina |  | 8848 | 1062 | 12% | 1947 | 22% |
|  |  |  |  | 1227 | 380 | 31% | 503 | 41% |
|  |  |  |  | 324 | 75 | 23% | 100 | 31% |

**Section 2. Additional data processing: Data aggregation, adjustments, and confidence intervals**

**2.1 Grouping conditions**

All 21 groups of conditions associated with SHS are further grouped into communicable diseases, non-communicable diseases, and injuries using the  categorization of IHME i.e.: 1) communicable diseases, which includes hemorrhagic fever, tuberculosis, HIV, and infectious diseases of the central neural system, 2) injuries, and 3) non-communicable diseases, which includes malignant neoplasms (except leukemia), leukemia, dementia, degenerative diseases of the central neuron system such as Parkinson’s diseases and multiple sclerosis, cerebrovascular diseases, chronic rheumatic heart diseases, cardiomyopathy and heart failure, chronic ischemic heart diseases, lung diseases, diseases of liver, renal failure, low birth weight and prematurity, birth trauma, congenital malformations, atherosclerosis, musculoskeletal disorders, protein-energy malnutrition, and endocrine, metabolic, blood and immune disorders. Unlike in IHME/GBD data, maternal, child and nutritional causes are grouped together with other non-communicable diseases for the purpose of analyzing SHS2.0.

**2.2 Double counting**

When calculating the total burden of SHS in non-decedents, double counting could be a major cause of over-estimation since country data aggregate numbers across all conditions. This is addressed in SHS2.0 for co-morbidities in non-decedent categories of four conditions identified as the most likely sources of double counting: Kaposi sarcoma in HIV patients, cancer, cerebrovascular diseases, and dementia. After analyzing the data, the following adjustments were undertaken: 1) Kaposi sarcoma patients are only counted for HIV but not for malignant neoplasms; 2) cancer patients with dementia and/or cerebrovascular disease are counted for cancer but not for the other two conditions; and 3) among non-cancer patients, those with cerebrovascular disease and dementia are counted for cerebrovascular disease but not for dementia. Details of the algorithm for these adjustments are presented in Appendix Panel 1.

**2.3 Confidence intervals**

Another improvement in SHS 2.0 is the use of relevant data from the IHME/GBD database to generate confidence intervals. Confidence intervals were constructed based on the IHME data for each of the 21 health conditions considered according to mortality and prevalence in the corresponding GBD codes for each year. For this, based on the mean values and the limits of variation reported by the IHME itself, 1,000 iterations of assuming a normal distribution around the mean and with standard deviation equal to
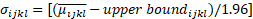
where
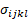
is the standard deviation with respect to the mean of condition *i*, in country *j*, in age group *k*, and sex *l* were carried out. Likewise,
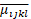
corresponds to the estimated mean value of SHS based on the data reported by IHME in the GBD data for condition *i*, in country *j*, in age group *k*, and sex *l*; while
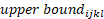
represents the upper limit of SHS; and 1.96 corresponds to the z-score for a 95% confidence interval in a standard normal distribution. This process was stratified by year, health condition, country, age group, and sex. Following this, SHS multipliers were applied to either the prevalence of mortality in each iteration to derive a specific SHS value. The mean of these values across the 1,000 iterations was considered the expected value. The 95% confidence interval was then defined by the range from the 2.5th to the 97.5th percentiles of these values.

**Appendix Panel 1:** Details on two adjustments to remove overlap between conditions and related double counting

**Removing Kaposi Sarcoma (KS) from cancer survivors experiencing SHS**

Almost all KS patients are also PLWHIVs, and they can account for up to 26% of total cancer survivors in some countries, causing a considerable overestimation of the number of patients in need of palliative care. Since KS patients’ experiences of suffering are more closely aligned with typical HIV patients than cancer patients, KS patients are removed from the cancer non-decedents.

Country-specific KS prevalence data are available from GLOBOCAN IARC cancer registries (note that GBD does not separately estimate KS). The 2018 data are used since only the most recent GLOBOCAN data include KS prevalence. Country-specific total cancer prevalence can also be downloaded from GLOBOCAN for consistency reasons. A KS % is calculated for each country: *KS % = country-specific KS prevalence / country-specific total cancer prevalence*. This percentage is then used to adjust the cancer non-decedents in need of PC for each country such that: *Cancer non-decedents_after adjustment = Cancer non-decedents_before adjustment * (1-KS%)*. No KS adjustment is made to the years 1990, 2000 and 2010 due to data availability limitations.

**Removing overlap among cancer, dementia and stroke survivors experiencing SHS**

Cancer, dementia, and stroke are selected among all non-decedent categories to adjust for overlaps because: 1) they share patients with similar age profiles; and 2) there is a large number of total global survivors thus the overestimation caused by double counting could pose as serious issue.

The following two assumptions are applied: 1) the prevalence of each disease is independent of each other, meaning that a cancer patient has the same possibility of living with dementia as the general population; 2) the suffering hierarchy is as follows: cancer > cerebrovascular disease > dementia, meaning that people with more than one condition will only be counted under the one condition causing the greatest suffering. Prevalence rate is calculated for each country, which is downloaded from the IHME database. The recommended formula. to remove double counting among cancer, cerebrovascular disease and dementia patients is as follows:

|  | **Cancer (C)** | **cerebrovascular disease (V)** | **Dementia (D)** |
| --- | --- | --- | --- |
| **Non-decedents category include:** | C, C-D, C-V, C-D-V | V, V-D, C-V, C-D-V | D, C-D, V-D, C-D-V |
| **Non-decedents category include_adjusted:** | C, C-D, C-V, C-D-V | V, V-D | D |
| **Adjustment** | No changes | Pv*(1-Pc) | Pd*(1-Pc)*(1-Pv) |
